# Social isolation and mortality risk in late-stage older Japanese: A longitudinal study of community-dwelling residents from 2020 to 2024

**DOI:** 10.1101/2024.12.13.24318347

**Authors:** Yukari Okawa

## Abstract

**Objectives:** To examine the association between social isolation and mortality among older adults aged ≥75 years in Japan.

**Study design:** A retrospective longitudinal study included homebound Japanese adults aged ≥75 years who participated in voluntary health checkups conducted by Zentsuji City, Kagawa Prefecture, Japan, between 2000 and 2024.

**Methods:** The relationship between social isolation and mortality was assessed using the Cox proportional hazards model. Social isolation was defined as the absence of regular contact with family or friends, and survival information was confirmed in the city’s database as of 1 July 2024.

**Results:** Of the 3366 participants (male: 42.4%), 3024 (male: 42.6%) remained in the final cohort. At study entry, socially isolated participants tended to be older, less likely to exercise, have worse self-rated health and life satisfaction, and have no family members in the same household. The mean follow-up time was 2.47 years, during which 9.1% of the participants died. Mortality risk was 2.37 times higher (95% confidence interval [CI]: 1.61–3.48) for socially isolated individuals than for those who were not socially isolated, after controlling for covariates such as physical status, self-rated health, lifestyle, life satisfaction, and sharing a household with family members. The results showed a consistent trend, even after excluding participants with short follow-up periods.

**Conclusion:** Based on data from older adults in Japan, a population with the world’s longest life expectancy, the findings of this study may help guide public health interventions aimed at reducing social isolation in today’s globally aging society.

## Introduction

Humans are social beings. Social determinants that influence a higher risk of death include employment status, educational level, marital status, and type of health insurance^1^. Having fewer social relationships is also known to be a risk factor for all-cause mortality^2^. Social relationship assessments include both structural (e.g., social networks, living alone, social isolation, and marital status) and functional (e.g., receiving and perceiving social support) measures. Importantly, social isolation is a known risk factor for higher mortality^2^.

Social isolation, which affects 25.0% of the world population, has historically been identified by several factors, including having less interaction with others, being unmarried, and not belonging to a social group^3–5^. A meta-analysis of 1.3 million middle-aged and older populations worldwide reported that social isolation carries a 1.33-fold (95% confidence interval [CI]: 1.26–1.41) higher risk of death than the absence of social isolation^6^. Furthermore, social isolation affects more people in North America (14 studies; hazard ratio [HR] 1.41 [95% CI: 1.28–1.55]) and Europe (10 studies; HR 1.33 [95% CI: 1.20–1.47]) than in Asia (5 studies; HR 1.20 [95% CI: 1.12–1.27])^6^. Three of five previous cohort studies in Asia targeted the Japanese population, with a relatively young baseline age (range: mid-60s to early 70s)^5,7,8^.

Japan has the world’s longest-living society. One-third of the population in 2023 was over 65 years old, and life expectancy at birth in 2021 was 84.5 years, which is the longest of all 38 Organization for Economic Cooperation and Development (OECD) members, including Asian countries such as South Korea (83.6 years), China (78.1 years), India (70.2 years), and Indonesia (68.8 years)^9,10^. Furthermore, a study of older adults reported that those in Japan are more socially isolated (8.7%) than those in Britain (1.3%), and social isolation has a more significant impact on mortality among Japanese people (adjusted hazard ratio [aHR] 1.18 [95% CI: 1.05–1.33]) than British people (aHR 1.27 [95% CI: 0.85–1.89])^7^. Furthermore, socially isolated, homebound older Japanese people were at a 2.19 times higher risk of dying than their non-socially isolated, non-homebound peers^5^. This suggests that more research is required to assess the relationship between social isolation and subsequent death in older, homebound Japanese populations.

Therefore, this longitudinal study evaluated the relationship between social isolation and subsequent death in the late-stage older homebound Japanese population.

## Methods

### Study design, data source, and study population

This retrospective longitudinal study included homebound Japanese citizens of Zentsuji City, Kagawa Prefecture, Japan, who underwent annual health checkups (2020–2024)^11,12^. The checkup targeted Japanese individuals aged 75 years or older at fiscal year (FY) age. All data were extracted from the city’s database on 1 July 2024^12^. In total, 5575 city residents were ≥75 at FY age (male: 39.6%) on 1 December 2023^12^. In FY2023, a total of 41.0% of those eligible for checkups underwent voluntary checkups.

The content of the annual checkups followed the protocol of the Japanese Ministry of Health, Labour and Welfare^11^. Annual checkups include anthropometric and blood pressure measurements, a blood test, urinalysis, and a self-reported questionnaire asking about the examinee’s social isolation, lifestyle, cognitive function, and their awareness of their physical and mental health.

### Social isolation

In this analysis, the time-fixed exposure variable was the participants’ social isolation. Participants who responded “no” to the question, “Do you regularly interact with family and friends? (yes/no),” were classified as socially isolated.

### Death

The outcome variable was defined as death occurring after the participant’s final annual checkup. On 1 July 2024, participants’ survival status (alive/dead) was confirmed via the city’s database; however, the dates of death were unavailable^12^. Therefore, the last date of the FY of the last visit to the annual checkup was treated as the date of death in this study.

In Japan, the FY begins on 1 April and ends on 31 March of the following year. Therefore, 31 March 2022 was, for example, given as the approximated date of death for a participant whose last recorded visit was during FY2021, and whose survival information was identified as “deceased” on 1 July 2024. For FY2024 only, 1 July 2024—the confirmation date for survival data—was treated as the end of the FY.

### Covariates

The following information was used in our analysis as potential confounding factors: biological sex (male[reference]/female); age (74–79[reference]/80–89/≥90); overweight or obese (no[reference]/yes); gamma-glutamyl transferase (GGT) quartiles (Q1[reference]/Q2/Q3/Q4); self-reported smoking status (non- or ex-smoker[reference]/smoker); self-reported exercise habits such as walking at least once a week (self-reported exercise status) (no[reference]/yes); self-rated health (good or somewhat good[reference]/normal/not very good or not good); hypertension (no[reference]/yes); dyslipidemia (no[reference]/yes); hemoglobin A1C (HbA1c); estimated glomerular filtration rate (eGFR); self-reported daily life satisfaction (satisfied or somewhat satisfied[reference]/unsatisfied or somewhat unsatisfied); having family members in the same household (yes[reference]/no); and residential district (East[reference]/Tatsukawa/South/Fudeoka/Central/West/Yoshiwara/Yogita)^5–8,13^.

Overweight or obese was defined as body mass index (BMI) ≥25.0 kg/m^214^. Because information about alcohol intake was unavailable in this study, GGT quartiles were used as an indicator of heavy drinking^15^. The GGT quartiles for males and females, in order, were as follows: Q1 = <17 and <14 U/L; Q2 = 17–23 and 14–16 U/L; Q3 = 24–36 and 17–23 U/L; and Q4 = ≥37 and ≥24 U/L.

Hypertension was defined as systolic blood pressure of ≥130 mmHg and/or diastolic blood pressure ≥80 mmHg^16^. Dyslipidemia was regarded as serum low-density lipoprotein cholesterol of ≥140 mg/dL, serum high-density lipoprotein cholesterol of <40 mg/dL, and/or serum triglycerides of ≥150 mg/dL^17^. HbA1c (%) was reported in the National Glycohemoglobin Standardization program units^18^. The eGFR (mL/min/1.73m^2^) was calculated using the three-variable equation for Japanese: 194 × serum creatinine in mg/dL^-1.094^ × age in years^-0.287^ (× 0.739, if female)^19^. The presence of family members in the same household was confirmed according to the city’s tax household records; therefore, whether the family actually lives together was not verified.

### Statistical analysis

To evaluate the relationship between social isolation and subsequent death in the older adult Japanese population, participants were defined as “Japanese aged ≥75 years old in FY” and “for whom data on the exposure and outcome variables were available.” **Figure 1** illustrates the exclusion process of participants. The length of time at risk for each participant was expressed in person-years, calculated from the date of the first checkup to the last date of the FY of the last visit. One exception was for those whose last visit was during FY2024: the date of confirmation of the participant’s survival, 1 July 2024, was used as the last date of FY2024.

**Fig. 1.**
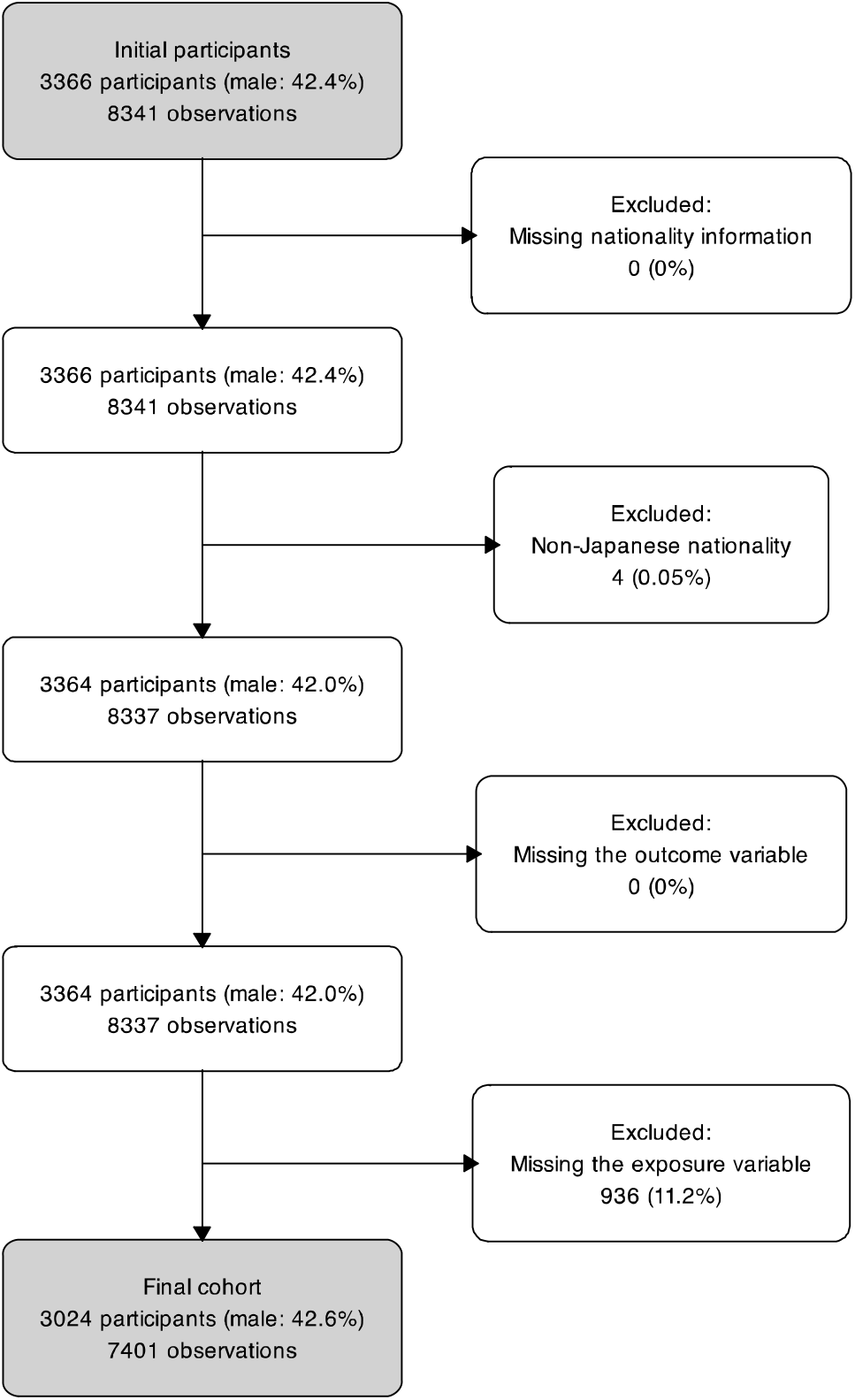
Participants flow diagram

The baseline characteristics of participants were summarized, stratified by the exposure variable, with continuous variables presented as median (interquartile range [IQR]) and categorical variables as numbers and proportions. Then, a map was created showing the eight districts of Zentsuji City and their respective proportions of social isolation. Kaplan–Meier curves were also created, stratified by the exposure variable. The proportional hazards (PH) assumption was tested, and no violations were found^20^.

Owing to the nature of voluntary checkups, the data contain missing values (% missing of the total observations): overweight or obesity (1.0%); GGT (1.0%); self-reported smoking status (0.1%); self-reported exercise status (0.4%); self-rated health (0.1%); hypertension (1.0%); dyslipidemia (1.0%); HbA1c (1.0%); eGFR (3.1%); self-reported daily life satisfaction (0.2%); and residential district (0.5%). Missing values were imputed by the multiple imputation method using chained equations (continuous variables by linear regression, binary variables by logistic regression, ordinal variables by ordered logistic regression, and nominal variables by multinomial logistic regression)^21^. The number of imputations was set to 20^22^.

After the imputations, the relationship between the time-fixed exposure (no social isolation[reference]/social isolation) and subsequent death was assessed using the Cox PH model, with the Efron method for tied failure and with the clustered sandwich estimator by each participant to adjust for standard errors caused by repeated measures. HRs and 95% CIs were used as an outcome index to compare survival between the exposure groups.

In addition to constructing a crude model, we adjusted Model 1 for sex and age category; Model 2 was further adjusted for overweight or obesity, GGT quartiles, self-reported smoking status, self-reported exercise status, self-rated health, hypertension, dyslipidemia, HbA1c, and eGFR; Model 3 was further adjusted for self-reported daily life satisfaction; and Model 4 was further adjusted for family members in the same household and the residential districts. To stabilize the models, a multiplicative term was added to the adjusted models when we observed an interaction between the exposure variable and covariates (sex, age category, overweight or obesity, GGT quartiles, self-reported smoking status, self-reported exercise status, self-rated health, hypertension, dyslipidemia, HbA1c, eGFR, family members in the same household, and the residential district).

Throughout, we treated a two-sided p-value of <0.05 as statistically significant and followed the Strengthening the Reporting of Observational Studies in Epidemiology (STROBE) for reporting observational studies. All statistical analyses were conducted using Stata/MP 16.1 (StataCorp, College Station, TX, USA). The city map and the flowchart showing the exclusion process were drawn using R version 4.4.2^9^.

### Sensitivity analyses

Two sensitivity analyses were conducted in this study. In the first sensitivity analysis, participants who underwent a checkup only once and were later confirmed to have died were excluded to minimize the impact of those who were close to death. The second sensitivity analysis used the generalized gamma (GG) model to examine how the results of the PH model differed from those of the flexible parametric accelerated failure-time (AFT) model. In the GG model, time ratios (TRs) and 95% CIs were used as an outcome index (e.g., TR <1 means shorter survival than the reference).

## Results

### Main analysis

The number of initial participants was 3366 (male: 42.4%), with a median age at baseline (IQR) of 78 (75–83) years. After exclusions, 3024 participants (male: 42.6%) remained in the final cohort, with a median age at baseline (IQR) of 79 (76–84) years (**Figure 1**). The total and mean follow-up times were 7460.3 person-years and 2.47 years, respectively. Of the participants in the total follow-up period, 4.9% were in the social isolation group at baseline. The mean follow-up time was shorter in the social isolation group (1.45 years) than in the group without social isolation (2.43 years). After the follow-up period, 9.1% of the total participants died: 31.8% in the social isolation group and 7.9% in the group without social isolation. **Figure 2** shows a city map with the proportions of socially isolated residents for each district.

**Fig. 2.**
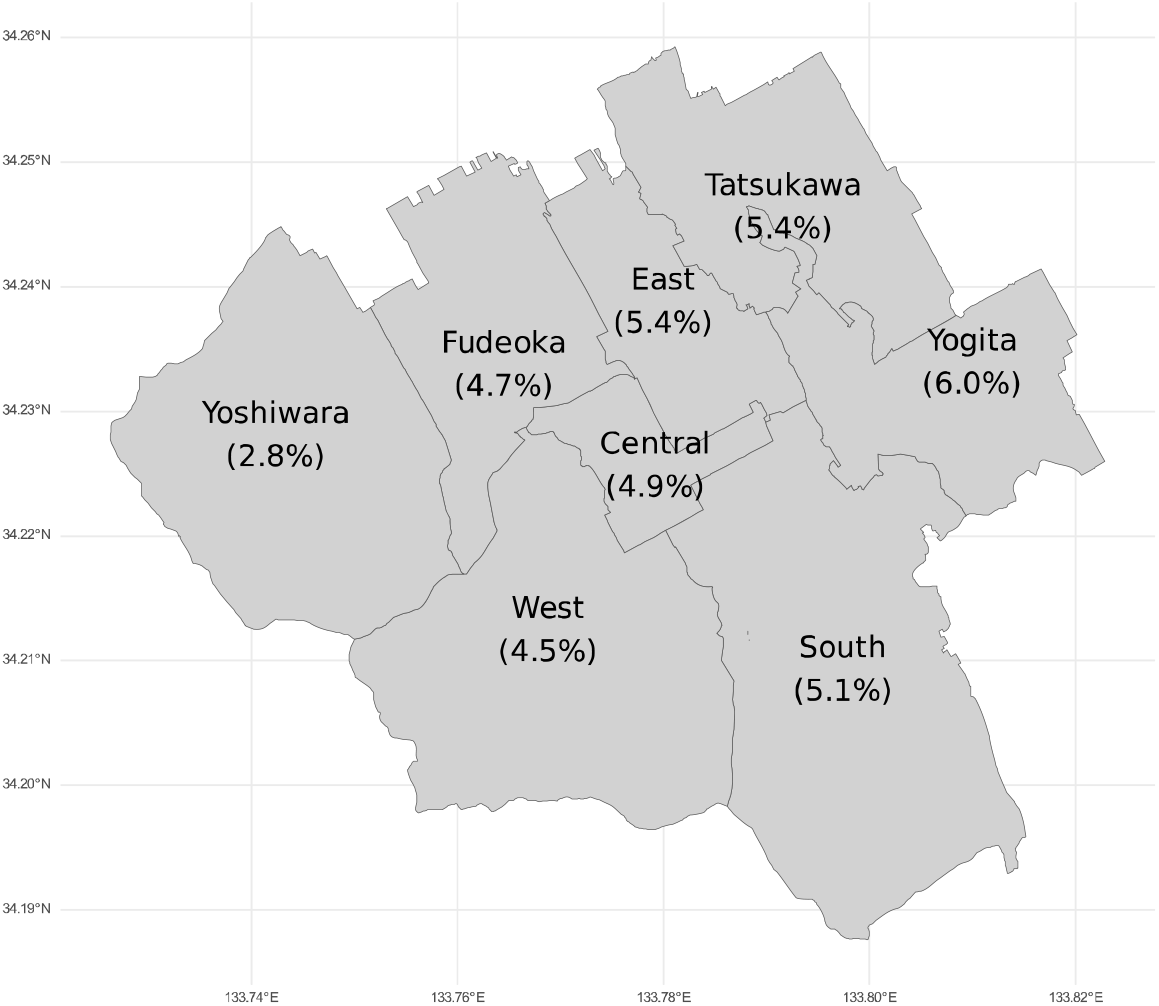
Map of Zentsuji City showing proportion of socially isolated residents in each district (%)

**Table 1** shows the participants’ characteristics at baseline, categorized by social isolation. Approximately 5% of participants belonged to the group with social isolation at baseline. Compared with participants who were not socially isolated at baseline, socially isolated participants at baseline tended to be older, have fewer exercise habits, worse self-rated health status, worse daily life satisfaction, and no family members in the same household.

**Table 1.**
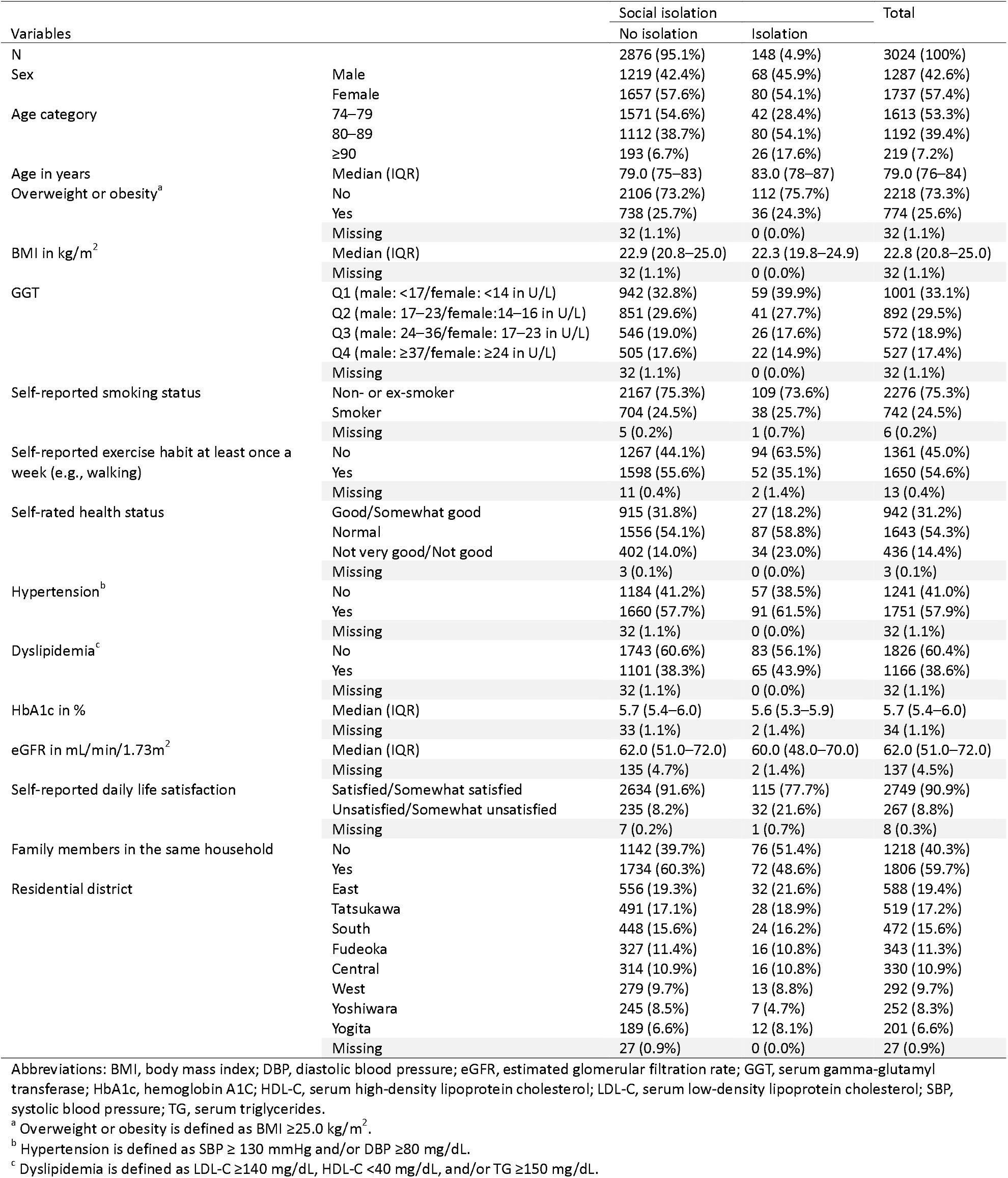
Baseline characteristics of study participants, stratified by social isolation.

| Variables |  | Social isolation |  | Total |
| --- | --- | --- | --- | --- |
|  |  | No isolation | Isolation |  |
| N |  | 2876 (95.1%) | 148 (4.9%) | 3024 (100%) |
| Sex | Male | 1219 (42.4%) | 68 (45.9%) | 1287 (42.6%) |
|  | Female | 1657 (57.6%) | 80 (54.1%) | 1737 (57.4%) |
| Age category | 74–79 | 1571 (54.6%) | 42 (28.4%) | 1613 (53.3%) |
|  | 80–89 | 1112 (38.7%) | 80 (54.1%) | 1192 (39.4%) |
|  | ≥90 | 193 (6.7%) | 26 (17.6%) | 219 (7.2%) |
| Age in years | Median (IQR) | 79.0 (75–83) | 83.0 (78–87) | 79.0 (76–84) |
| Overweight or obesity<sup>a</sup> | No | 2106 (73.2%) | 112 (75.7%) | 2218 (73.3%) |
|  | Yes | 738 (25.7%) | 36 (24.3%) | 774 (25.6%) |
|  | Missing | 32 (1.1%) | 0 (0.0%) | 32 (1.1%) |
| BMI in kg/m<sup>2</sup> | Median (IQR) | 22.9 (20.8–25.0) | 22.3 (19.8–24.9) | 22.8 (20.8–25.0) |
|  | Missing | 32 (1.1%) | 0 (0.0%) | 32 (1.1%) |
| GGT | Q1 (male: <17/female: <14 in U/L) | 942 (32.8%) | 59 (39.9%) | 1001 (33.1%) |
|  | Q2 (male: 17–23/female: 14–16 in U/L) | 851 (29.6%) | 41 (27.7%) | 892 (29.5%) |
|  | Q3 (male: 24–36/female: 17–23 in U/L) | 546 (19.0%) | 26 (17.6%) | 572 (18.9%) |
|  | Q4 (male: ≥37/female: ≥24 in U/L) | 505 (17.6%) | 22 (14.9%) | 527 (17.4%) |
|  | Missing | 32 (1.1%) | 0 (0.0%) | 32 (1.1%) |
| Self-reported smoking status | Non- or ex-smoker | 2167 (75.3%) | 109 (73.6%) | 2276 (75.3%) |
|  | Smoker | 704 (24.5%) | 38 (25.7%) | 742 (24.5%) |
|  | Missing | 5 (0.2%) | 1 (0.7%) | 6 (0.2%) |
| Self-reported exercise habit at least once a week (e.g., walking) | No | 1267 (44.1%) | 94 (63.5%) | 1361 (45.0%) |
|  | Yes | 1598 (55.6%) | 52 (35.1%) | 1650 (54.6%) |
|  | Missing | 11 (0.4%) | 2 (1.4%) | 13 (0.4%) |
| Self-rated health status | Good/Somewhat good | 915 (31.8%) | 27 (18.2%) | 942 (31.2%) |
|  | Normal | 1556 (54.1%) | 87 (58.8%) | 1643 (54.3%) |
|  | Not very good/Not good | 402 (14.0%) | 34 (23.0%) | 436 (14.4%) |
|  | Missing | 3 (0.1%) | 0 (0.0%) | 3 (0.1%) |
| Hypertension<sup>b</sup> | No | 1184 (41.2%) | 57 (38.5%) | 1241 (41.0%) |
|  | Yes | 1660 (57.7%) | 91 (61.5%) | 1751 (57.9%) |
|  | Missing | 32 (1.1%) | 0 (0.0%) | 32 (1.1%) |
| Dyslipidemia<sup>c</sup> | No | 1743 (60.6%) | 83 (56.1%) | 1826 (60.4%) |
|  | Yes | 1101 (38.3%) | 65 (43.9%) | 1166 (38.6%) |
|  | Missing | 32 (1.1%) | 0 (0.0%) | 32 (1.1%) |
| HbA1c in % | Median (IQR) | 5.7 (5.4–6.0) | 5.6 (5.3–5.9) | 5.7 (5.4–6.0) |
|  | Missing | 33 (1.1%) | 2 (1.4%) | 34 (1.1%) |
| eGFR in mL/min/1.73m<sup>2</sup> | Median (IQR) | 62.0 (51.0–72.0) | 60.0 (48.0–70.0) | 62.0 (51.0–72.0) |
|  | Missing | 135 (4.7%) | 2 (1.4%) | 137 (4.5%) |
| Self-reported daily life satisfaction | Satisfied/Somewhat satisfied | 2634 (91.6%) | 115 (77.7%) | 2749 (90.9%) |
|  | Unsatisfied/Somewhat unsatisfied | 235 (8.2%) | 32 (21.6%) | 267 (8.8%) |
|  | Missing | 7 (0.2%) | 1 (0.7%) | 8 (0.3%) |
| Family members in the same household | No | 1142 (39.7%) | 76 (51.4%) | 1218 (40.3%) |
|  | Yes | 1734 (60.3%) | 72 (48.6%) | 1806 (59.7%) |
| Residential district | East | 556 (19.3%) | 32 (21.6%) | 588 (19.4%) |
|  | Tatsukawa | 491 (17.1%) | 28 (18.9%) | 519 (17.2%) |
|  | South | 448 (15.6%) | 24 (16.2%) | 472 (15.6%) |
|  | Fudeoka | 327 (11.4%) | 16 (10.8%) | 343 (11.3%) |
|  | Central | 314 (10.9%) | 16 (10.8%) | 330 (10.9%) |
|  | West | 279 (9.7%) | 13 (8.8%) | 292 (9.7%) |
|  | Yoshiwara | 245 (8.5%) | 7 (4.7%) | 252 (8.3%) |
|  | Yogita | 189 (6.6%) | 12 (8.1%) | 201 (6.6%) |
|  | Missing | 27 (0.9%) | 0 (0.0%) | 27 (0.9%) |

Abbreviations: BMI, body mass index; DBP, diastolic blood pressure; eGFR, estimated glomerular filtration rate; GGT, serum gamma-glutamyl transferase; HbA1c, hemoglobin A1C; HDL-C, serum high-density lipoprotein cholesterol; LDL-C, serum low-density lipoprotein cholesterol; SBP, systolic blood pressure; TG, serum triglycerides.
<sup>a</sup> Overweight or obesity is defined as BMI ≥25.0 kg/m<sup>2</sup>.
<sup>b</sup> Hypertension is defined as SBP ≥ 130 mmHg and/or DBP ≥80 mg/dL.
<sup>c</sup> Dyslipidemia is defined as LDL-C ≥140 mg/dL, HDL-C <40 mg/dL, and/or TG ≥150 mg/dL.

Figure 3. presents Kaplan–Meier curves and categorizes the number of older people at risk of death by social isolation (log-rank p<0.001). The estimation results for the Cox PH model are presented in **Table 2**. The incidence rate was higher in the group with social isolation. Socially isolated participants had a higher mortality risk—2.37 times (95% CI: 1.61–3.48)—than those who were not socially isolated, even after adjusting for all covariates. When the analysis was conducted by sex, the crude HRs (95% CIs) were higher for females at 3.22 (2.09–4.95) than for males at 2.06 (1.30–3.25). The models adjusted for sex did not fit well, probably owing to the small number of participants (male: n=1309, mean follow-up time of 2.38 years; female: n=1746, mean follow-up time of 2.49 years) and high-dimensional data^23^.

**Table 2.** Social isolation and subsequent death among 3024 older Japanese citizens of Zentsuji City, Kagawa Prefecture, Japan (7401 observations, 2020–2024)^a^.

| Variable | Follow-up information |  |  | Models |  |  |  |  |
| --- | --- | --- | --- | --- | --- | --- | --- | --- |
|  | Total time at risk (PY) | Failures (n) | IR (rates per 1000 PY) | Crude HR (95% CI) | Model 1 aHR (95% CI) | Model 2 aHR (95% CI) | Model 3 aHR (95% CI) | Model 4 aHR (95% CI) |
| Social isolation |  |  |  |  |  |  |  |  |
| No isolation (reference) | 7118.4 | 227 | 31.9 | 1.00 | 1.00 | 1.00 | 1.00 | 1.00 |
| Isolation | 341.9 | 47 | 137.5 | 2.61 (1.92–3.54) | 2.37 (1.73–3.26) | 2.01 (1.44–2.81) | 1.94 (1.36–2.76) | 2.37 (1.61–3.48) |

Abbreviations: aHR, adjusted hazard ratio; BMI, body mass index; DBP, diastolic blood pressure; eGFR, estimated glomerular filtration rate; GGT, serum gamma-glutamyl transferase; HbA1c, hemoglobin A1C; HR, hazard ratio; HDL-C, serum high-density lipoprotein cholesterol; IR, incidence rate; LDL-C, serum low-density lipoprotein cholesterol; PY, person-years; SBP, systolic blood pressure; TG, serum triglycerides.
<sup>a</sup> The Cox proportional hazards model was conducted.
Multiple imputed variables: ALT, AST, dyslipidemia<sup>d</sup>, eGFR, GGT, HbA1c, hypertension<sup>c</sup>, overweight or obesity<sup>b</sup>, residential district, and self-reported smoking status.
<sup>b</sup> Overweight or obesity is defined as BMI $\geq 25.0$ kg/m<sup>2</sup>.
<sup>c</sup> Hypertension is defined as SBP $\geq 130$ mmHg and/or DBP $\geq 80$ mg/dL.
<sup>d</sup> Dyslipidemia is defined as LDL-C $\geq 140$ mg/dL, HDL-C $< 40$ mg/dL, and/or TG $\geq 150$ mg/dL.
Model 1: Adjusted for sex (male[reference]/female) and age category (74–79[reference]/80–89/ $\geq 90$ ).
Model 2: Adjusted for both variables of Model 1, overweight or obesity (no[reference]/yes)<sup>b</sup>, GGT quartiles (Q1[reference]/Q2/Q3/Q4), self-reported smoking status (non- or ex-smoker[reference]/smoker), self-reported exercise habit at least once a week (yes[reference]/no), and self-rated health (Good or somewhat good[reference]/normal/not very good or not good), hypertension (no[reference]/yes)<sup>c</sup>, dyslipidemia (no[reference]/yes)<sup>d</sup>, HbA1c, and eGFR.
Model 3: Adjusted for all variables of Model 2 and self-reported daily life satisfaction (satisfied or somewhat satisfied[reference]/unsatisfied or somewhat unsatisfied).
Model 4: Adjusted for all variables of Model 3, family members in the same household (yes[reference]/no), and the residential district
(East[reference]/Tatsukawa/South/Fudeoka/Central/West/Yoshiwara/Yogita). A multiplicative term (social isolation × self-reported smoking status) was added.

### Sensitivity analyses

#### First sensitivity analysis: exclusion of participants with short follow-up

**Table S1** shows the results of the first sensitivity analysis, which assessed the potential influence of participants nearing death by excluding those who passed away after the first observation. After exclusions, the total follow-up time was only reduced by 1.0% (4.1% for the socially isolated group and 0.9% for the group that was not socially isolated).

**Fig. 3.**
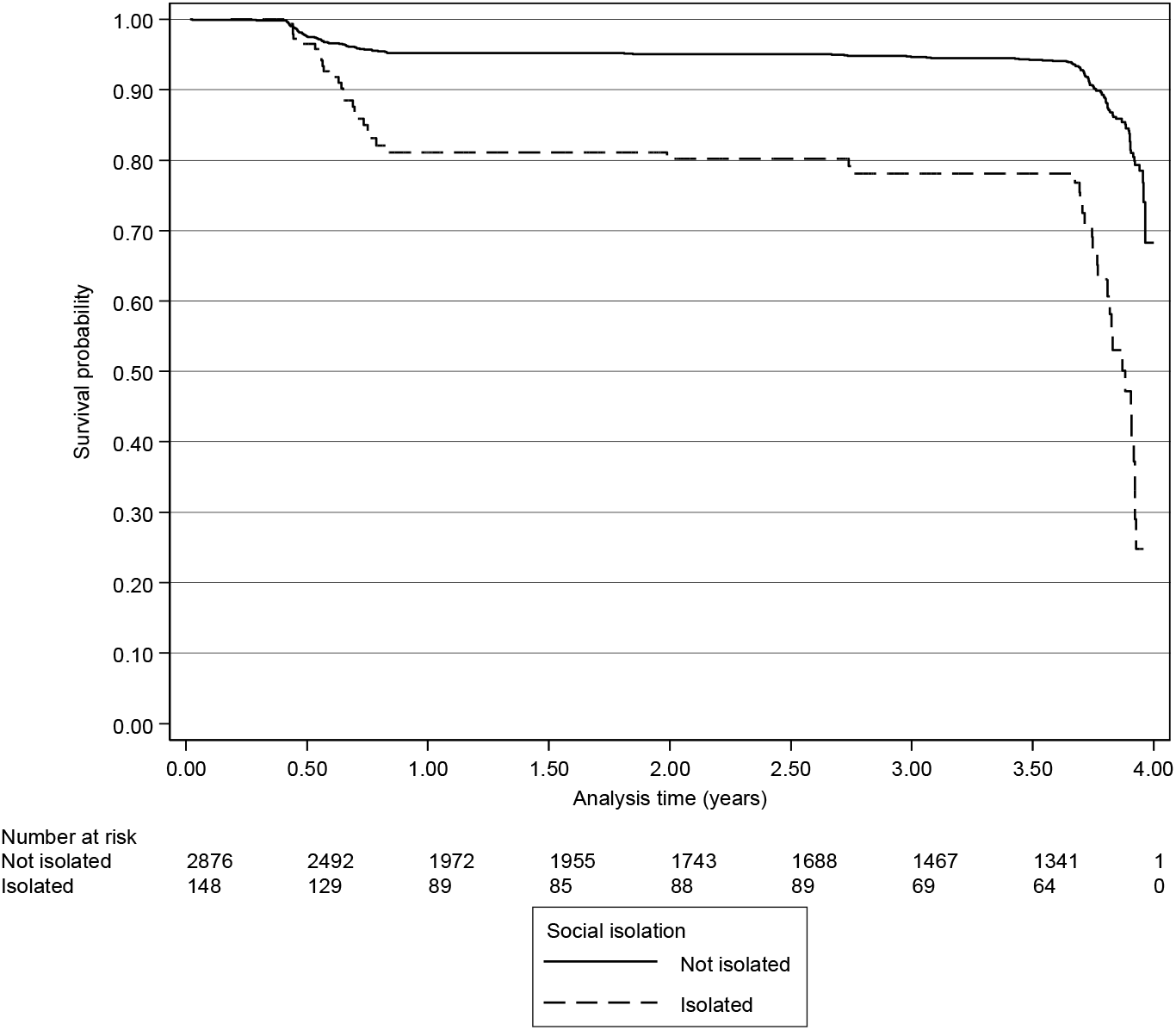
Kaplan–Meier curves and table showing mortality risk for older people, stratified by social isolation

The median age (IQR) at death was 89.5 (85–92) years for people who were socially isolated and 86 (83–92) for those who were not, which was similar to the results from the main analysis. Compared with the main analysis (**Table 2**), the results were slightly attenuated but similar: social isolation was a mortality risk, with an aHR of 2.18 (95% CI: 1.38–3.44). This implies no significant effect of the inclusion of participants at high risk of death.

#### Second sensitivity analysis: parametric AFT model

In the second sensitivity analysis, using the parametric AFT model with the GG distribution reduced the survival time of socially isolated people by 4% (95% CI: 1%–7%) compared with those who were not socially isolated (**Table S2**). The tendency for social isolation to increase the risk of death was unchanged from the results of the main analysis (**Table 2**). This similar trend also implies no explicit effect of time-varying covariates.

## Discussion

This longitudinal study of homebound Japanese citizens aged ≥75 years shows that social isolation is a risk factor for death among older adults, after controlling for effects from possible covariates, including sex, age, physical status, lifestyle, and even life satisfaction and the presence of family members in the same household. Overall, the findings of the present study support those of previous studies of older Japanese adults^7,8^; however, we observed a higher point estimate and wider 95% CIs than those in the other studies. More specifically, the Aichi Gerontological Evaluation Study (AGES) reported an aHR of 1.19 (95% CI: 1.02–1.39) and the Japan Gerontological Evaluation Study (JAGES) reported an aHR of 1.18 (95% CI: 1.05–1.33)^7,8^.

Several factors may account for this difference. First, our participants were predominantly homebound and older than those in the previous studies, which introduces potential selection bias^7,8^. A previous cohort study of adults aged ≥65 years at baseline reported that mortality risk was higher among homebound people (aHR 2.19 [95% CI: 1.04–4.63]) than non-homebound people (aHR 1.34 [95% CI: 0.64– 2.81]) if they were socially isolated, compared with non-homebound people who were not socially isolated^5^. Furthermore, older age is an independent risk factor for death. Therefore, the point estimates of this study were higher than those reported in the AGES and JAGES, possibly owing to the difference in the target population (i.e., the late-stage older people living at home)^7,8^.

Second, the difference may be partly explained by how social isolation is defined. Because this study was a secondary data analysis, data on social isolation as assessed by the commonly used scales, such as the Berkman-Syme Social Network Index (SNI) and the Lubben Social Network Scale (LSNS), were unavailable^24–26^. Indeed, in each of the mentioned studies, social isolation was defined differently^7,8^. Participants in this study were simply asked if they regularly interact with their friends and family (yes/no), whereas participants in the AGES were asked about the frequency of face-to-face or non-face-to-face (letter, telephone, and email) contacts with family members and relatives per month (6 responses), and in the JAGES participants reported face-to-face and non-face-to-face contacts with children, relatives, and friends (7 responses). Because each study’s definition differed from the others, the proportions of social isolation at baseline also vary: 4.9% (no regular contact) in the current study, 15.5% (≤2 contacts per month) in the AGES, 8.7% (<1 contact per month) in the JAGES, and 5.5% (score of ≥3) in the JAGES social isolated risk score (**Table 1**). In this study, some socially isolated people were classified as non-isolated because the question limited regular interaction to “friends and family,” and the frequency and depth of interactions that were considered “regular” depended on respondents’ perceptions^11^. This misclassification may have led to an underestimation of the association.

Third, because this study used secondary data, some important factors were unavailable^11^. For example, socioeconomic status (e.g., higher income, married) is a confounding factor that reduces the risk of social isolation and mortality^7,27–29^. Without adjusting for these variables, all estimates are likely to be overestimated. To mitigate the effect of marital status and living alone on the results, this study instead adjusted for having family members in the same household, which yielded consistent results.

Finally, this study included only 3024 older residents of Zentsuji City, which had a population of 30,336 as of 1 December 2023^12^. Thus, the small study population may have induced greater variability in estimates, resulting in fluctuating point estimates and wider 95% CIs.

### Strengths and Limitations

The main strength of this study is its focus on evaluating the association between social isolation and death among relatively healthy older Japanese residents living independently in the community rather than in hospitals or nursing homes. The findings offer guidance for interventions among older populations to prevent social isolation and extend healthy life expectancies.

Several limitations should also be stated. The first and most significant limitation of this study was the unavailability of date-of-death information. We could confirm participants’ survival information using the city’s database as of 1 July 2024, but deceased participants had to have died between their last checkup and the point at which survival information was verified. Therefore, to avoid inappropriately prolonged survival times for those who had died or had no subsequent earlier visit during the follow-up period, the end of the FY of the last visit was treated as the date of death or censored.

The second limitation was the use of all-cause mortality as the outcome variable. All-cause mortality includes deaths that are unrelated to social isolation, such as accidental death. However, in 2023, accidental deaths by proportion of total deaths by age group in Japan were 2.98% for people in their 60s, 2.93% in their 70s, 2.91% in their 80s, 2.32% in their 90s, and 1.69% for those aged ≥100^9^. Therefore, because the proportions of deaths attributed to accidental causes were low among older respondents, it is likely that the use of all-cause mortality as the outcome variable in this study did not produce an overestimation of the results.

The third limitation was the short follow-up time, from 2020 to 2024 (mean follow-up time: 2.47 years)^11^. Because the risk of death increases with time, a <5-year follow-up time may not capture the event within the observation period; therefore, it underestimates the association between social isolation and death. However, the results of the first sensitivity analysis, which excluded participants who died after the first observation, showed a similar trend to the main analysis (**Tables 2 and S1**). Thus, it is unlikely that a short follow-up period substantially influenced the results.

The fourth limitation was the possible persistence of residual bias related to age, such as factors associated with a high risk of mortality and social isolation (e.g., cognitive decline, activities of daily living [ADL] impairment), which may have potentially influenced the findings, even after controlling for age^30–32^. However, people with severe illness who are unable to communicate with others or who have serious ADL problems tend not to get voluntary checkups on their own. Therefore, the effect of this bias may be smaller than that for the general Japanese population.

The fifth limitation was that physician-diagnosed mental disorders (e.g., depression) were not measured in this analysis. Without adjusting for mental disorders, the relationship between social isolation and mortality may be overestimated because depression is a known cause of social isolation and increased risk of mortality^30,33^. Interestingly, a study conducted in South Korea reported that patients with depression aged ≥65 years were 2.65 times (95% CI: 1.69–4.16) less likely to engage in health-seeking behaviors than those without depression^34^. This suggests that the impact of depression on the findings of this study may be limited, as people with depression may be less likely to attend checkups.

## Conclusions

In this longitudinal study (2020–2024) of 3024 older Japanese adults living in the community, socially isolated people had a higher risk of death than those who were not, after adjusting for all covariates. The findings indicate that older people who do not interact regularly with anyone may hasten their death, even if they are physically healthy and have their families nearby. Further research should not only aim to prolong lives, but also seek specific interventions, such as encouraging socialization, that can improve happiness and health across the lifespan.

## Acknowledgments

The author expresses gratitude to all participants in this study, as well as to Ayaka Nakatsu, Masako Matsumoto, Mayumi Kitadani, and the local government officers of Zentsuji City for their invaluable support. We thank Anita Harman, PhD, from Edanz (https://jp.edanz.com/ac) for editing a draft of this manuscript.

## Author statements

### Ethical approval

The data were anonymized before receipt. The Ethics Committee of Okayama University Graduate School of Medicine, Dentistry and Pharmaceutical Sciences and Okayama University Hospital approved this study (No. K1708-040). The ethics committee waived the need for informed consent. This research followed the Declaration of Helsinki and Japanese Ethical Guidelines for Medical and Biological Research involving Human Subjects.

### Data availability statement

All data generated or analyzed during this study are available within the main text of the published article and its supplementary information files.

