## Supplementary material for "Social isolation and mortality risk in late-stage older Japanese: A longitudinal study of community-dwelling residents from 2020 to 2024"

**Table S1**. Social isolation and subsequent death among 2881 older Japanese citizens of Zentsuji City, Kagawa Prefecture, Japan, excluding those who died after the first observation (7258 observations, 2020–2024)[a].

| | Follow-up information | | | Models | | | | |
|---|---|---|---|---|---|---|---|---|
| | Total time at risk | Failures | IR | Crude | Model 1 | Model 2 | Model 3 | Model 4 |
| Variable | (PY) | (n) | (rates per 1000 PY) | HR (95% CI) | aHR (95% CI) | aHR (95% CI) | aHR (95% CI) | aHR (95% CI) |
| Social isolation | | | | | | | | |
| No isolation (reference) | 7055.3 | 107 | 15.2 | 1.00 | 1.00 | 1.00 | 1.00 | 1.00 |
| Isolation | 327.8 | 24 | 73.2 | 2.69 (1.83–3.97) | 2.31 (1.53–3.49) | 2.18 (1.40–3.40) | 2.12 (1.33–3.38) | 2.18 (1.38–3.44) |

Abbreviations: aHR, adjusted hazard ratio; BMI, body mass index; DBP, diastolic blood pressure; eGFR, estimated glomerular filtration rate; GGT, serum gamma-glutamyl transferase; HbA1c, hemoglobin A1C; HR, hazard ratio; HDL-C, serum high-density lipoprotein cholesterol; IR, incidence rate; LDL-C, serum low-density lipoprotein cholesterol; PY, person-years; SBP, systolic blood pressure; TG, serum triglycerides.

[a] The Cox proportional hazards model was conducted.

Multiple imputed variables: ALT, AST, dyslipidemia[d], eGFR, GGT, HbA1c, hypertension[c], overweight or obesity[b], residential district, and self-reported smoking status.

[b] Overweight or obesity is defined as BMI ≥25.0 kg/m$^2$.

[c] Hypertension is defined as SBP ≥ 130 mmHg and/or DBP ≥80 mg/dL.

[d] Dyslipidemia is defined as LDL-C ≥140 mg/dL, HDL-C <40 mg/dL, and/or TG ≥150 mg/dL.

Model 1: Adjusted for sex (male[reference]/female) and age category (74–79[reference]/80–89/≥90).

Model 2: Adjusted for both variables of Model 1, overweight or obesity (no[reference]/yes)[b], GGT quartiles (Q1[reference]/Q2/Q3/Q4), self-reported smoking status (non- or ex-smoker[reference]/smoker), self-reported exercise habit at least once a week (yes[reference]/no), and self-rated health (Good or somewhat good[reference]/normal/not very good or not good), hypertension (no[reference]/yes)[c], dyslipidemia (no[reference]/yes)[d], HbA1c, and eGFR.

Model 3: Adjusted for all variables of Model 2 and self-reported daily life satisfaction (satisfied or somewhat satisfied[reference]/unsatisfied or somewhat unsatisfied).

Model 4: Adjusted for all variables of Model 3, family members in the same household (yes[reference]/no), and the residential district (East[reference]/Tatsukawa/South/Fudeoka/Central/West/Yoshiwara/Yogita).

**Table S2**. Social isolation and subsequent death among 3024 older Japanese citizens of Zentsuji City, Kagawa Prefecture, Japan: the generalized gamma model (7401 observations, 2020–2024).

| | Follow-up information | | | Models | | | | |
|---|---|---|---|---|---|---|---|---|
| | Total time at risk | Failures | IR | Crude | Model 1 | Model 2 | Model 3 | Model 4 |
| Variable | (PY) | (n) | (rates per 1000 PY) | TR (95% CI) | aTR (95% CI) | aTR (95% CI) | aTR (95% CI) | aTR (95% CI) |
| Social isolation | | | | | | | | |
| No isolation (reference) | 7118.4 | 227 | 31.9 | 1.00 | 1.00 | 1.00 | 1.00 | 1.00 |
| Isolation | 341.9 | 47 | 137.5 | 0.93 (0.90–0.97) | 0.95 (0.92–0.97) | 0.96 (0.93–0.98) | 0.96 (0.93–0.99) | 0.96 (0.93–0.99) |

Abbreviations: aHR, adjusted hazard ratio; BMI, body mass index; DBP, diastolic blood pressure; eGFR, estimated glomerular filtration rate; GGT, serum gamma-glutamyl transferase; HbA1c, hemoglobin A1C; HR, hazard ratio; HDL-C, serum high-density lipoprotein cholesterol; IR, incidence rate; LDL-C, serum low-density lipoprotein cholesterol; PY, person-years; SBP, systolic blood pressure; TG, serum triglycerides.

Multiple imputed variables: ALT, AST, dyslipidemia[c], eGFR, GGT, HbA1c, hypertension[b], overweight or obesity[a], residential district, and self-reported smoking status.

[a] Overweight or obesity is defined as BMI ≥25.0 $kg/m^2$.

[b] Hypertension is defined as SBP ≥ 130 mmHg and/or DBP ≥80 mg/dL.

[c] Dyslipidemia is defined as LDL-C ≥140 mg/dL, HDL-C <40 mg/dL, and/or TG ≥150 mg/dL.

Model 1: Adjusted for sex (male[reference]/female) and age category (74–79[reference]/80–89/≥90).

Model 2: Adjusted for both variables of Model 1, overweight or obesity (no[reference]/yes)[a], GGT quartiles (Q1[reference]/Q2/Q3/Q4), self-reported smoking status (non- or ex-smoker[reference]/smoker), self-reported exercise habit at least once a week (yes[reference]/no), and self-rated health (Good or somewhat good[reference]/normal/not very good or not good), hypertension (no[reference]/yes)[b], dyslipidemia (no[reference]/yes)[c], HbA1c, and eGFR.

Model 3: Adjusted for all variables of Model 2 and self-reported daily life satisfaction (satisfied or somewhat satisfied[reference]/unsatisfied or somewhat unsatisfied).

Model 4: Adjusted for all variables of Model 3, family members in the same household (yes[reference]/no), and the residential district (East[reference]/Tatsukawa/South/Fudeoka/Central/West/Yoshiwara/Yogita).
